# Oscillations in USA COVID-19 Incidence and Mortality Data reflect societal factors

**DOI:** 10.1101/2020.06.08.20123786

**Authors:** Aviv Bergman, Yehonatan Sella, Peter Agre, Arturo Casadevall

**Author notes:** Address Correspondence Arturo Casadevall Johns Hopkins School of Public Health 615 N. Wolfe Street Room E5132 Baltimore, Maryland 21205.

## Abstract

The COVID-19 pandemic currently in process differs from other infectious disease calamities that have previously plagued humanity in the vast amount of information that is produces each day, which includes daily estimates of the disease incidence and mortality data. Apart from providing actionable information to public health authorities on the trend of the pandemic, the daily incidence reflects the process of disease in a susceptible population and thus reflects the pathogenesis of COVID-19, the public health response and diagnosis and reporting. Both daily new cases and daily mortality data in the US exhibit periodic oscillatory patterns. By analyzing NYC and LA testing data, we demonstrate that this oscillation in the number of cases can be strongly explained by the daily variation in testing. This seems to rule out alternative hypotheses such as increased infections on certain days of the week as driving this oscillation. Similarly, we show that the apparent oscillation in mortality in the US data is mostly an artifact of reporting, which disappears in datasets that record death by episode date, such as the NYC and LA datasets. Periodic oscillations in COVID-19 incidence and mortality data reflect testing and reporting practices and contingencies. Thus, these contingencies should be considered first prior to suggesting social or biological mechanisms.

---

Epidemic case incidence data provides raw information on the course and outcome of infectious disease outbreaks. Such data informs on whether the epidemic is surging, ebbing or evolving into an endemic. Trends in the case incidence data provide insight into the efficacy of medical and behavioral interventions, such as vaccines and social distancing measures, respectively. In addition, trends in case incidence data as a function of time reflect biological parameters of the host-microbe interaction including prevalence of disease among infected individuals, contagiousness of infection, diagnostic capacity and the size of the susceptible population. Since the case incidence data must ultimately be gathered, compiled and analyzed, the numbers also reflect the public health capacity and infrastructure available to track the course of the epidemic. Consequently, the case incidence data is a number that reflects many variables ranging from biological constraints of the host and pathogen entities to medical, diagnostic, public health and social structures on the affected population.

Since case incidence data reflect the conglomerations of many different informational components with variable contributions it is difficult to tease out their individual contributions from trend curves. Given the complexity of how epidemic case incidence data emerges, epidemiological modeling is a necessary and important tool for understanding how individual variables contribute to this number. However, it is possible that higher order analysis of case incidence data produces patterns that reflect the contribution of the individual contributory components. For example, an analysis of case incidence data in Germany revealed an oscillatory pattern that was attributed to weekly fluctuations in case reporting (1). Consequently, we analyzed the trends COVID-19 case incidence data in the USA to see if we could discern patterns that provided additional insight into the course of the epidemic.

## Methods

USA daily new case and death data was obtained from https://github.com/nytimes/covid-19-data/blob/master/us.csv for 111 days, from January 21^st^ until May 11^th^. In addition, data from New York City was obtained from https://github.com/nychealth/coronavirus-data, including total daily tests as well as daily positive tests (Figure 1A,B), and deaths (Figure 2B). Similarly, LA data was obtained from http://dashboard.publichealth.lacounty.gov/covid19_surveillance_dashboard/. Initially, a power spectrum analysis on the raw data was performed using MATLAB (https://www.mathworks.com/help/matlab/ref/rand.html) Fast Fourier Transform Analysis (2). We then performed a 7-day rolling average on each time series in order to smooth the data, and then divided the original time series by its smoothed version in order to obtain a normalized time series which measures deviation from the smoothed data, thus capturing the short-term variations in the data while controlling for the longer-term trend. We then performed regression and correlation analyses on the normalized time series, comparing the normalized daily positive tests to normalized daily total tests.

**Figure 1:**
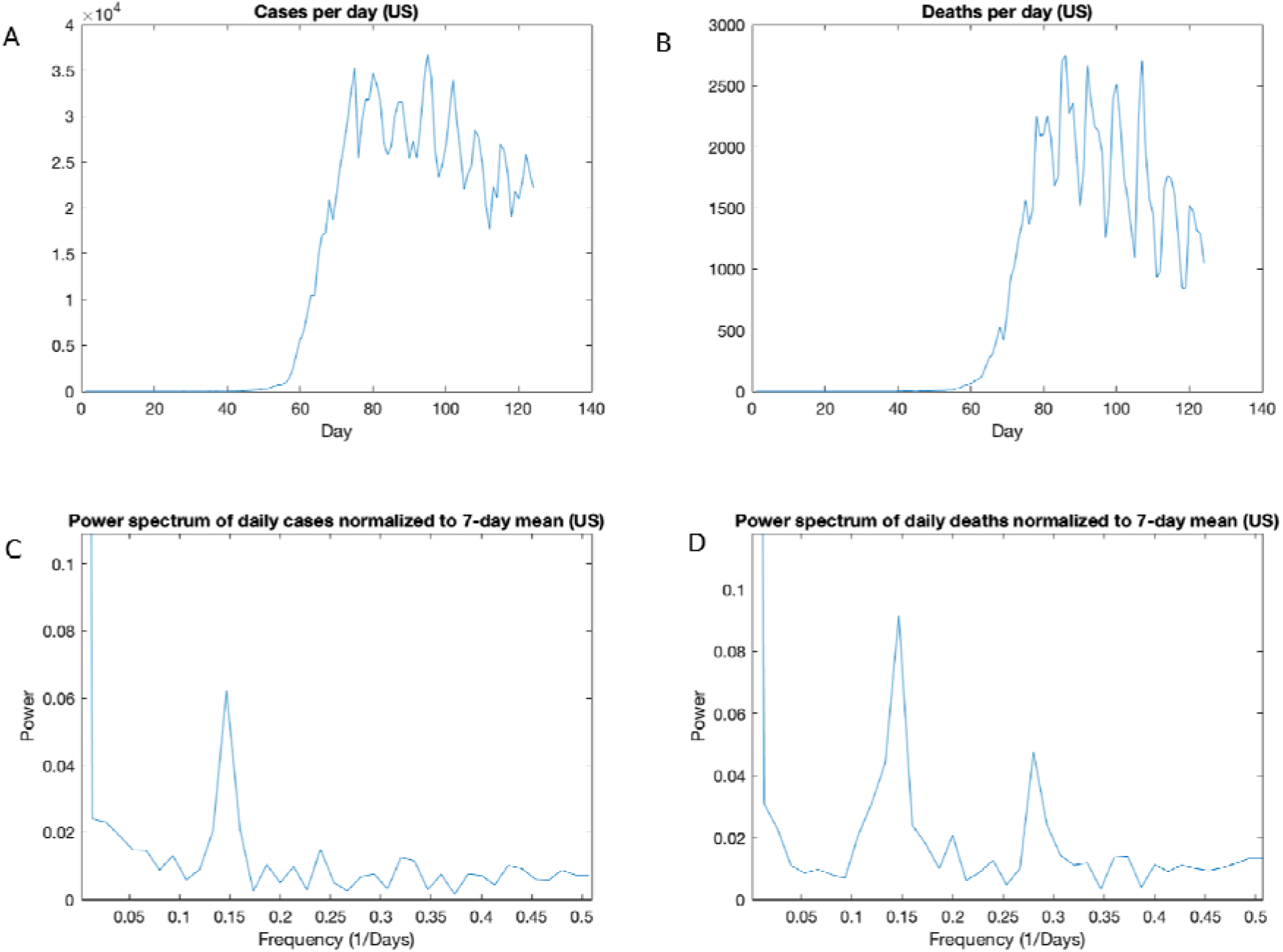
Periodic behavior in COVID-19 cases and deaths in the US. A) Daily cases in the US time. B) Daily deaths in the US over time. C) Power spectrum of daily cases in the US,normalizalized to a 7-day moving average. D) Power spectrum of daily deaths in the US, normalize to a 7-day moving average.

**Figure 2:**
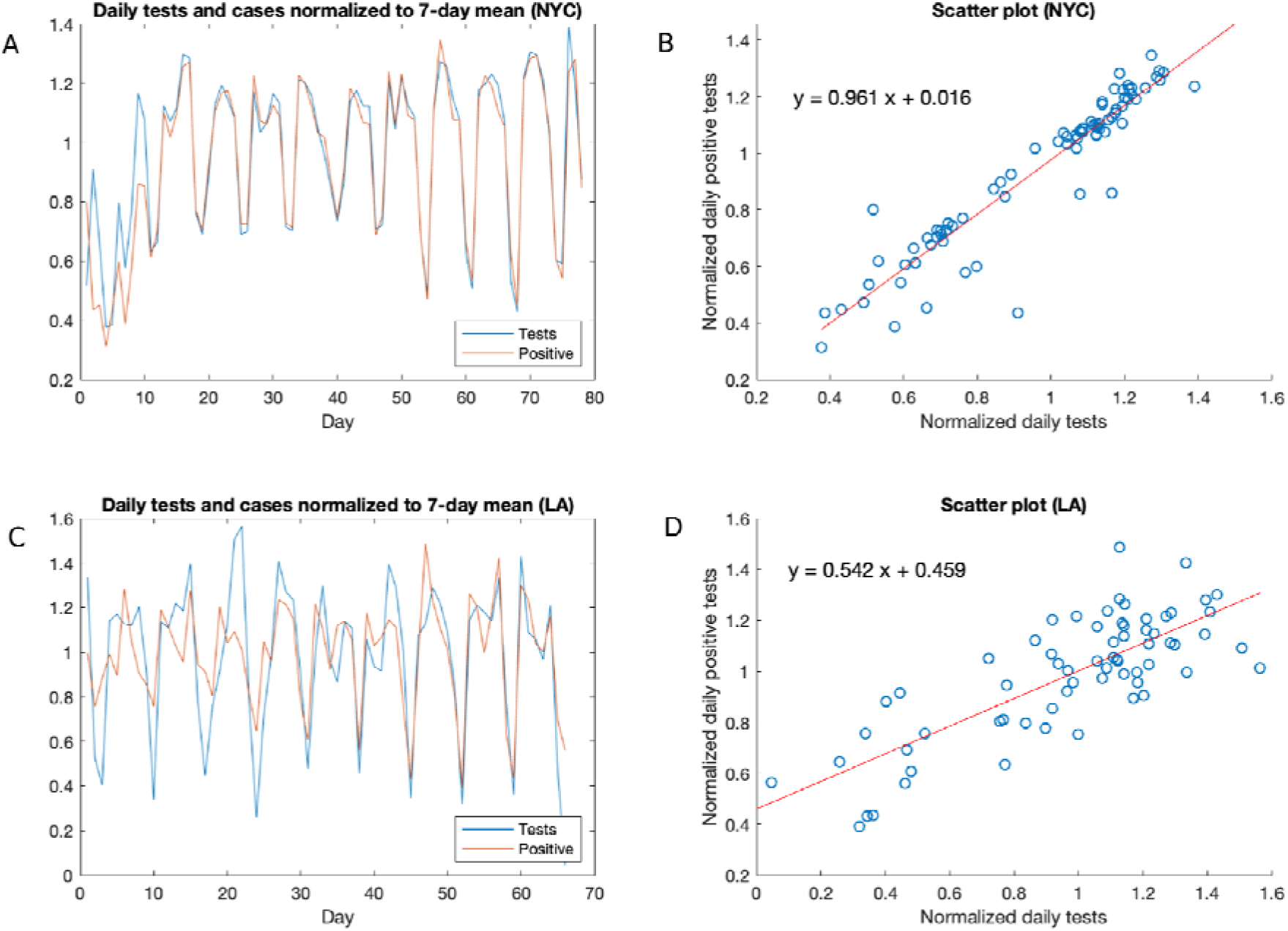
Analysis of daily cases and deaths in NYC and LA. A) Daily tests and positive tests in NYC over time, normalized to a 7-day moving average. B)Scatter plot and regression line for the normalized tests and positive tests data in NYC. C)Daily tests and positive tests in LA over time, normalized to a 7-day moving average. D)Scatter plot and regression line for the normalized tests and positive tests data in LA.

## Results and Discussion

To quantify the observed oscillation in incidence and mortality, we performed a power spectrum analysis of US cases, which revealed a clear period corresponding to every 6.8 days (Figure 1C). A similar analysis of NYC cases revealed a major period of every 7.1 days, and minor periods of every 3.6 and 2.4 days. The presence of clear gaps between peaks in the power spectrum signifies periodic patterns as opposed to chaotic or stochastic signatures (3, 4). Mortality data in the US also revealed a periodicity of 6.8 days (Figure 1D).

The periodicities of 6.8 and 7.1 days are very close to the length of a week, suggesting societal rather than purely biological factors are at play. The more minor periodicities of 3.6 and 2.4 could potentially arise artificially due to their nature as approximately ½ and 1/3 of a week. This weekly periodicity could have more than one explanation. One trivial explanation is that it represents an increase in cases related to diagnostic and reporting activity related to the tempo of the week. For example, greater reporting of cases on some days of the week as backlogs are cleared from weekend testing could produce an increases in reported cases that, repeated each week, will create a recurring periodicity. To investigate the hypothesis that the oscillations in incidence were simply a reflection of the testing done we focused on the NYC and LA datasets, where testing and incidence data were available, and which also have the advantage of being localized regions with high incidence. The number of tests in NYC has a clear valley on Sundays and peaks on Wednesdays-Thursdays, and the number of reported positive follows. Indeed, when both total tests and positive tests are normalized to a 7 day average, we observe a near perfect correlation between the two (r = 0.94) (Figure 2A). Moreover, the two normalized time series align almost perfectly as seen visually (Figure 2B) or by the fact that the line y = x fits the scatter plot well (as seen in Figure 1D, it is close to the line of best fit). Data in LA yields similar, though less strong, results, probably due to lower number of cases giving rise to more noise (Figure 2C,D). This suggests that alternative potential explanations, such as increased social activity on certain days of the week leading to a greater infection rate, are not significant drivers of the observed oscillations.

Similarly, the oscillations in mortality data appear to be an artifact of reporting. We note an important difference between the US data and the datasets of NYC and LA we have obtained: while in the US dataset, case and death dates reflect reporting date, the NYC and LA data is back-dated to the episode date. The NYC and LA datasets are therefore more reliable indicators of mortality, and indeed we do not observe a weekly periodicity in NYC deaths. When normalized to a 7-day moving average, the NYC and LA mortality data show some oscillation, but with no clear periodicity (Figure 3C,D). Indeed, the power spectra of the normalized mortality data do not yield clear peaks (Figure 3E,F), indicating a stochastic rather than periodic signal.

**Figure 3:**
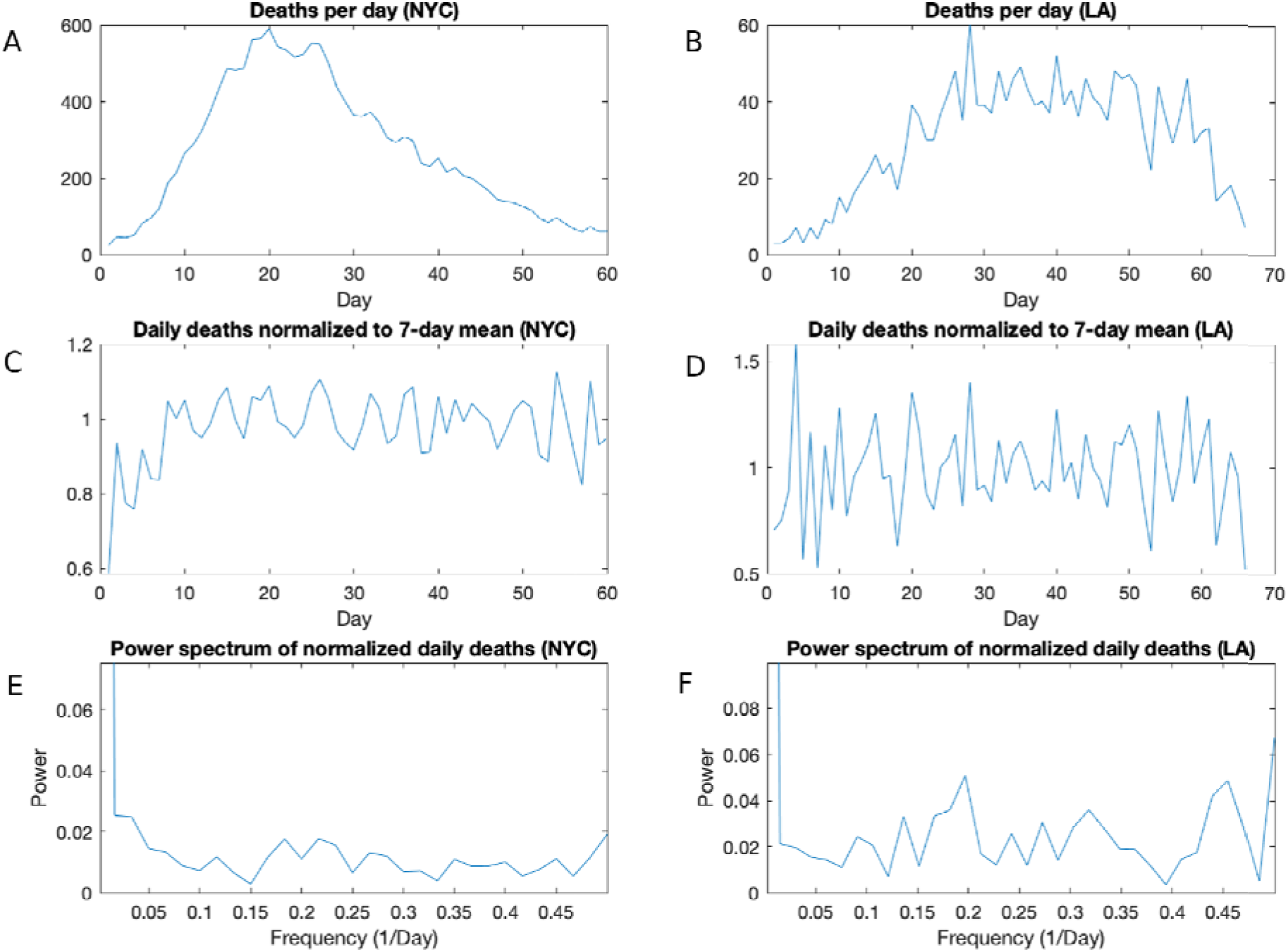
Analysis of mortality data in NYC and LA. A) Daily deaths in NYC; B) daily deaths in LA; c)Daily deaths in NYC, normalized to a 7-day moving average; D) Daily deaths in LA, normalized to a 7-day moving average; E) Power spectrum of the normalized mortality data in NYC, revealing no no clear period. F) Power spectrum of daily deaths in LA, revealing no clear period.

Several studies have shown variations in medical care between weekdays and weekends. In this regard, patients having procedures later in the week and in weekends had higher mortality than those treated earlier in the week (5). Similarly, individuals admitted to hospitals during weekends have higher mortality than those admitted during the week (6). However, such variations in medical care do not appear to drive clear periodicity in NYC and LA mortality data.

Regardless of the above caveats, we considered the possibility of biological causes for oscillation but were able to rule these out. The periodicity at 2.4 days observed in NYC incidence data is very close to the time required between infection and capacity to spread the virus, which was estimated to be 2.3 d (7), bringing up the question of whether this more minor periodicity reflects any biological process. However, such a time delay is will not lead to periodicity in incidence unless it is coupled with periodic bursts of increased interaction and infection, for example, as may be caused by increased activity in the weekend. However, our above analysis of NYC data, demonstrating a strong correlation with testing, ruled out increased infection rate as a significant driver of oscillation, making it unlikely that the time delay of 2.3 days manifests in periodicity in the data. Indeed, the strong correlation of incidence with testing data suggests that the periodicity of 2.3 days does not reflect real periodicity in incidence. This reasoning led us away from any biological explanation of the apparent periodicities.

In summary, we observed periodic weekly oscillations in the incidence and mortality data for COVID-19. The oscillations in incidence data showed a very strong correlation to the day-today testing numbers implying that these were directly caused by variation in testing. Mortality oscillations appear to reflect the weekly tempo of reporting, rather than true oscillations in deaths. Hence, oscillations of incidence and mortality data could point to biasing practices in case reporting which should be considered first prior to suggesting social or biological mechanisms..

## Data Availability

Data available to all who request it

## References

1. Hotz T, Glock M, Heyder S, Semper S, Böhle A, Krämer A. Monitoring the spread of COVID-19 by estimating reproduction numbers over time. arXiv preprint arXiv:200408557. 2020.

2. Bracewell RN, Bracewell RN. The Fourier transform and its applications: McGraw-Hill New York; 1986.

3. Kulp CW. Detecting chaos in irregularly sampled time series. Chaos. 2013;23(3):033110. Epub 2013/10/05. doi: 10.1063/1.4813865. PubMed PMID: 24089946.

4. Kulp AWN, B.J. Characterization of time series data. In: Skiadas CHS. C., editor. Applications of Chaos Theory. Boca Raton, Fl: CRC Press; 2016. p. 131.

5. Aylin P, Alexandrescu R, Jen MH, Mayer EK, Bottle A. Day of week of procedure and 30 day mortality for elective surgery: retrospective analysis of hospital episode statistics. Bmj. 2013;346:f2424. Epub 2013/05/30. doi: 10.1136/bmj.f2424. PubMed PMID: 23716356; PMCID: Pmc3665889.

6. Bell CM, Redelmeier DA. Mortality among patients admitted to hospitals on weekends as compared with weekdays. The New England journal of medicine. 2001;345(9):663–8. Epub 2001/09/08. doi: 10.1056/NEJMsa003376. PubMed PMID: 11547721.

7. He X, Lau EHY, Wu P, Deng X, Wang J, Hao X, Lau YC, Wong JY, Guan Y, Tan X, Mo X, Chen Y, Liao B, Chen W, Hu F, Zhang Q, Zhong M, Wu Y, Zhao L, Zhang F, Cowling BJ, Li F, Leung GM. Temporal dynamics in viral shedding and transmissibility of COVID-19. Nature medicine. 2020;26(5):672–5. Epub 2020/04/17. doi: 10.1038/s41591-020-0869-5. PubMed PMID: 32296168.

